# Circulating Metabolites May Illustrate Relationship of Alcohol Consumption with Cardiovascular Disease

**DOI:** 10.1101/2023.05.24.23290487

**Authors:** Yi Li, Mengyao Wang, Xue Liu, Jian Rong, Patricia Emogene Miller, Roby Joehanes, Tianxiao Huan, Xiuqing Guo, Jerome Rotter, Jennifer Smith, Bing Yu, Matthew Nayor, Daniel Levy, Chunyu Liu, Jiantao Ma

## Abstract

**Background:** Metabolite signatures of long-term alcohol consumption are lacking. To better understand the molecular basis linking alcohol drinking and cardiovascular disease (CVD), we investigated circulating metabolites associated with long-term alcohol consumption and examined whether these metabolites were associated with incident CVD.

**Methods:** Cumulative average alcohol consumption (g/day) was derived from the total consumption of beer, wine and liquor on average of 19 years in 2,428 Framingham Heart Study Offspring participants (mean age 56 years, 52% women). We used linear mixed models to investigate the associations of alcohol consumption with 211 log-transformed plasma metabolites, adjusting for age, sex, batch, smoking, diet, physical activity, BMI, and familial relationship. Cox models were used to test the association of alcohol-related metabolite scores with fatal and nonfatal incident CVD (myocardial infarction, coronary heart disease, stroke, and heart failure).

**Results:** We identified 60 metabolites associated with cumulative average alcohol consumption (p<0.05/211≈0.00024). For example, one g/day increase of alcohol consumption was associated with higher levels of cholesteryl esters (e.g., CE 16:1, beta=0.023±0.002, p=6.3e-45) and phosphatidylcholine (e.g., PC 32:1, beta=0.021±0.002, p=3.1e-38). Survival analysis identified that 10 alcohol-associated metabolites were also associated with a differential CVD risk after adjusting for age, sex, and batch. Further, we built two alcohol consumption weighted metabolite scores using these 10 metabolites and showed that, with adjustment age, sex, batch, and common CVD risk factors, the two scores had comparable but opposite associations with incident CVD, hazard ratio 1.11(95% CI=[1.02, 1.21],p=0.02) vs 0.88 (95% CI=[0.78, 0.98], p=0.02).

**Summary:** We identified 60 long-term alcohol consumption-associated metabolites. The association analysis with incident CVD suggests a complex metabolic basis between alcohol consumption and CVD.

## INTRODUCTION

Alcohol drinking is a common lifestyle in many cultures and is a modifiable risk factor associated with over 200 health problems, including cardiovascular diseases (CVD) ^1^, dementia ^2^, neuropsychiatric conditions ^3^, liver cirrhosis,^4^ and diabetes^5^. For example, a systematic review and meta-analysis of 23 observational studies found that moderate alcohol consumption was related to a higher cardiovascular risk within 24 hours after alcohol intake; however, after 24 hours, moderate alcohol consumption seemed to have a protective effect on myocardial infraction ^6^. In contrast, heavy alcohol intake had a continued risk for cardiovascular events ^6^.

Metabolites are small molecules that are intermediates or end-products of metabolism in many cellular processes ^7, 8^. Metabolites can be quantified via high-throughput liquid chromatography with tandem mass spectrometry (LC/MS) methods.^9^ Association analyses of alcohol consumption and metabolites may help us gain insights into the effect of alcohol consumption on disease pathways. Several European studies investigated associations of total alcohol intake with circulating concentrations of metabolites^10–12^. One study investigated the association of 123 metabolites with alcohol intake using a discovery (n=1983) and replication (n=991) study design in healthy participants. This study found that 72 metabolites were significantly associated with alcohol consumption in the discovery set, and 34 metabolites remained significance in the replication set ^10^. A population-based study investigated 131 metabolites in 2090 individuals and revealed that 40 metabolites in men and 18 metabolites in women significantly differed in their concentrations between moderate-to-heavy and light alcohol drinking^12^.

While these metabolomic studies help us to understand the potential molecular basis of alcohol consumption, most studies analyzed the alcohol consumption measured at a single time point, which may not represent the habitual or long-term alcohol consumption. A better understanding of the relationships between long-term alcohol consumption and circulating metabolites, as well as the relationships of alcohol-associated metabolites with CVD risk, may help elucidate the complex relationship of alcohol consumption with etiology and progression of alcohol-associated diseases. To that end, our study used longitudinal data from the Framingham Heart Study (FHS) Offspring cohort to investigate the following three aims (Figure 1). First, we conducted association analyses of cumulative average alcohol consumption over 20 years with circulating metabolites. Second, we analyzed the specific associations of metabolites with cumulative consumption of three types of alcoholic beverages: beer, wine, and liquor. Third, we conducted association analyses of alcohol-associated metabolites with incident CVD.

**Figure 1.**
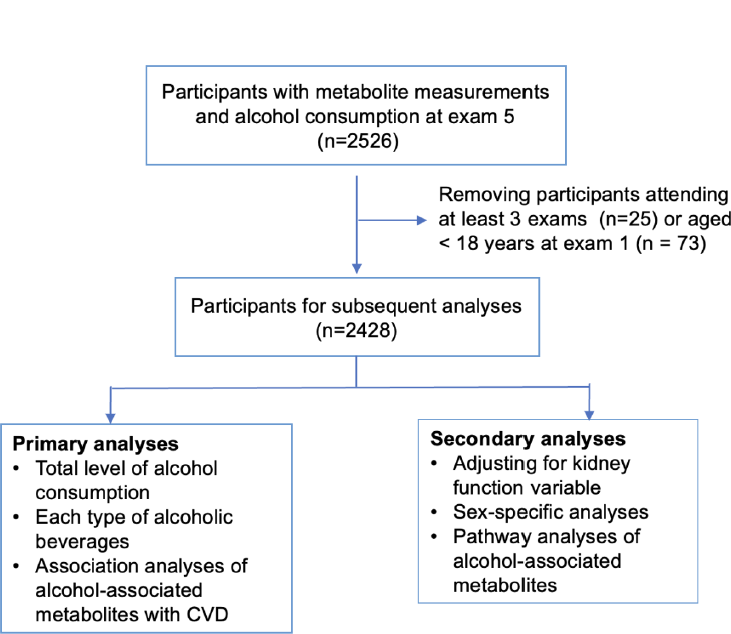
Flow-chart of Study.

## METHODS

### Population

The Offspring cohort (n = 5124) of the FHS was recruited in 1971, including the children of the Original cohort and spouses of these children^13^. The Offspring participants underwent in-person health examinations every four to eight years to collect comprehensive demographic and clinical measures such as risk factors for cardiovascular and neurodegenerative diseases^14^. This study analyzed Offspring participants who had metabolite measurements at the 5th examination (n=2526). We excluded participants who attended less than three examinations between examination 1 and examination 5 (n=25) and those who were younger than 18 years old at examination 1 (n=73). The final sample size was 2428 for statistical analyses (Figure 1). All participants provided written informed consent. All FHS study protocols were approved by the Boston University Medical Center Institutional Review Board.

### Metabolite quantification

Targeted metabolite profiling was conducted by LC/MS platform and hydrophilic interaction chromatography method as previously described^15–17^. The targeted metabolite profiling includes negatively charged polar metabolites (i.e., organic acids, bile acids, and sugars), positively charged polar metabolites (i.e., amino acids, urea cycle intermediates, nucleotides), and lipid metabolite species (i.e., cholesterol esters (CE), diacylglycerols (DAGs), lysophosphatidylcholines (LPCs), lysophosphatidylethanolamines (LPEs), phosphatidylcholines (PCs), sphingomyelins (SMs), triacylglycerols (TAGs)) ^15–18^. Two metabolites, TAG 54:5 and TAG 54:6, had identical value for all participants; we removed TAG 54:6 from the present analysis. Five metabolites were also excluded owing to large proportion of missingness in greater than 1000 participants, resulting in 211 metabolites for the subsequent analyses. Each metabolite measurement was natural log transformed, and then standardized to have mean at zero and standard deviation (SD) at 1. Metabolite values beyond ±4 SD away from the mean of that metabolite were set to be at ±4 SD ^17^.

### Alcohol consumption

At each examination, alcohol consumption information was collected through the FHS technician-administered questionnaires ^19^. The frequency of three types of alcoholic beverages (i.e., standard drinks of beer, wine, or liquor) in a typical week or month were collected. In this study, we used “grams of alcohol consumed per day”, which was derived from the summation the total standard drinks of beer, wine (red or white) or liquor. One standard drink was defined as one 12 oz. beer, one 4 oz. wine, or one 1.5 oz. 80 proof alcohol spirit, and one drink contains approximately 14 grams ethanol ^19, 20^. We calculated the grams per day for all types of alcoholic beverages (i.e., total alcohol consumption), as well as for each type (i.e., grams per day of beer, wine, or liquor). To reflect long-term alcohol consumption, we calculated the cumulative average alcohol consumption, i.e., the mean over up to? five examinations.

### Disease traits

CVD events were identified through adjudication by a panel of three physicians^21^. The CVD event data were obtained from annual health history updates based on inpatient or outpatient medical records, physical examinations, and mortality registry. CVD was comprised of myocardial infarction, coronary heart disease, stroke, heart failure, and death to any cardiovascular conditions^22^. We analyzed incident CVD outcome after removal of the prevalent cases at the 5^th^ examination.

### Covariates

Demographic information (sex and age), smoking status, dietary intake, time and intensity of physical activities, and medication use were obtained from standard questionnaire^23^. We calculated a physical activity index using the following formula: 1*sleep hours/day + 1.1*sedentary hours/day + 1.5*slight activity hours/day + 2.4*moderate activity hours/day + 5*heavy activity hours/day. We used Dietary Approaches to Stop Hypertension (DASH) score to define diet quality. We calculated the diet quality score based on consumptions of eight dietary components: vegetables, fruits, nuts and legumes, whole grains, dairy, red and processed meat, sugar-sweetened beverages and sodium^24^. Systolic blood pressure (SBP) was measured twice by physicians and the average blood pressure was used in the present analysis. Anthropometry was measured by technicians. Standard assays were used to determine levels of fasting glucose and plasma lipids. We defined diabetes if a participant had fasting glucose ≥126mg/dL or using diabetes medications.

### Statistical analysis

#### Association analysis of metabolites and alcohol consumption

In primary association analyses, we analyzed the relationships of metabolites (dependent variables) with cumulative average alcohol consumption of total alcohol, beer, wine, and liquor as independent variables (Figure 1). We used linear mixed models to quantify these associations, adjusting for age, sex, (metabolite measurement) batch, smoking status, BMI, physical activity, and diet quality score as fixed effect covariates (all measured at the 5th examination), and familial relationship as a random effect covariate. We used Bonferroni correction (p < 0.05/211≈0.00024) to account for multiple hypothesis testing with 211 metabolites.

We conducted two secondary analyses as described below. The level of alcohol consumption is usually larger in men compared to women ^25^, therefore, we tested sex-alcohol interaction by adding a production term of sex and alcohol consumption. We additionally adjusted for a serum creatinine-based estimated glomerular filtration rate (eGFR) to investigate if kidney function confounded the relationships between the cumulative average alcohol consumption and metabolites. (Figure 1).

#### Association analysis of alcohol-associated metabolites and CVD

Metabolites significantly associated with cumulative average alcohol consumption were used as independent variables in testing for associations with incident CVD. A Cox proportional hazards regression model was used, adjusting for sex, age and batch in the base model. In the multivariable model, we additionally adjusted for BMI, SBP, hypertension treatment status, diabetes, smoking status, total and high-density lipoprotein cholesterol levels. All covariates included were collected at the 5^th^ examination.

To study an aggregate effect of metabolites with the development of CVD, we constructed weighted composite scores with CVD-associated metabolites that were identified in the base model. We used estimates from the association analyses between cumulative average alcohol consumption and the metabolites to weigh the concentrations of the corresponding metabolites. We then calculated metabolite scores as the linear combination of weighted metabolite concentrations: *T_ij_* = ∑_*i*_ *W*_*i*_*V_ij_*. Two metabolite scores were calculated based on the direction of the associations between alcohol consumption, metabolites, and incident CVD. The first score aggregated metabolites with consistent direction, i.e., those were either positively or negatively associated with both alcohol consumption and CVD. The second score aggregated metabolites with opposite direction, i.e., those were positively associated with alcohol consumption and negatively associated with CVD or those were negatively associated with alcohol consumption and positively associated with CVD. The composite scores were standardized with mean at zero and standard deviation at one. We performed association analysis of the continuous composite scores with the development of CVD, adjusting for age, sex, batch, BMI, SBP, hypertension treatment status, diabetes, smoking status, total and high-density lipoprotein cholesterol levels. All association analyses were performed using R software (version 4.0.5).

#### Pathway analysis for metabolites

Pathway analyses were performed to identify biological pathways related to the alcohol-associated metabolites using MetaboAnalyst 5.0 ^26^. Two pathway libraries used in these analyses are Kyoto Encyclopaedia of Genes and Genomes (KEGG) and Small Molecule Pathway Database (SMPDB) for Homo sapiens. We conducted hypergeometric test with default parameters in MetaboAnalyst and reported significant pathways at false discovery rate (FDR) < 0.05.

## RESULTS

### Participant characteristics

This study included mostly middle-aged to older community dwelling men and women (n=2428, mean age=55.9 ± 9.3 years, 52% women at 5^th^ examination) (Table 1). The median of the cumulative average of total alcohol consumption was 7.7 g/day (interquartile range 16.8 g/day). Our participants were overweight (mean BMI 27.5 kg/m^2^) and 18.3% of them were current smokers at the 5th examination. Compared to women, men had greater alcohol consumption at all examinations: median consumption was 13.8 g/day in men versus 4.4 g/day in women at 5^th^ examination.

**Table 1.** Participants Characteristic.

| Variable | Male (N=1156) | Female (N=1272) | Total (N=2428) |
| --- | --- | --- | --- |
| Age, yrs | 56.5 (9.2) | 55.4 (9.4) | 55.9 (9.3) |
| Body mass index,kg/m<sup>2</sup> | 28.3 (4.1) | 26.9 (5.5) | 27.5 (4.9) |
| Physical activity index | 35.8 (7.2) | 33.3 (4.7) | 34.5 (6.2) |
| Diet score | 16.3 (4.3) | 17.9 (3.9) | 17.1 (4.1) |
| Estimated glomerular rate, ml/min/1.73m<sup>2</sup> | 73.8 (16.3) | 67.5 (14.7) | 70.5 (15.8) |
| Fasting blood glucose, mg/dL | 105.3 (31.7) | 97.9 (25.2) | 101.4 (28.7) |
| Total cholesterol, mg/dL | 203.0 (35.1) | 209.7 (37.9) | 206.5 (36.7) |
| High desity cholesterol, mg/dL | 42.9 (11.1) | 55.4 (15.4) | 49.4 (14.9) |
| Systolic blood pressure, mmHg | 129.7 (17.9) | 124.2 (19.6) | 126.9 (19.0) |
| Current smoker, n(%) | 208 (18.0%) | 236 (18.6%) | 444 (18.3%) |
| Cumulative average alcohol consumption variable, g/day; median (Q1, Q3) |  |  |  |
| Total alcohol consumption | 13.8 (5.3, 27.5) | 4.4 (1.7, 11.2) | 7.7 (2.5, 19.3) |
| Beer consumption | 4.4 (0.8, 13.4) | 0 (0, 0.8) | 0.8 (0, 5.2) |
| Wine consumption | 1.3 (0.4, 4.0) | 1.7 (0.4, 4.7) | 1.3 (0.4, 4.3) |
| Liquor consumption | 1.6 (0.4, 7.0) | 1.0 (0.4, 3.2) | 1.2 (0.4, 4.4) |
| Prevalent CVD | 114 (12.5%) | 78(6.1%) | 222(9.1%) |
| Incident CVD | 346(29.9%) | 290(22.8%) | 636(26.2%) |

The characteristics were measured at exam 5 (i.e., the baseline). Continuous variables are displayed as mean (standard deviation) or median (Q1-Q3). Binary variable is displayed as n (%)

### Associations of cumulative average alcohol consumption and metabolites

We found that 60 metabolites were significantly associated with the cumulative average alcohol consumption (p<0.05/211≈0.00024) adjusting for potential confounders (Table 2, **Supplemental Table 1**). Of the 60 metabolites, 40 metabolites displayed positive associations with the cumulative average alcohol consumption (**Supplemental Table 1**, Figure 2). The most significant metabolite was cholesteryl palmitoleate (CE16:1), a plasma cholesteryl ester that is involved in cholesterol metabolism ^27^. One g/day higher alcohol consumption was associated with higher level of cholesteryl palmitoleate (beta=0.023, p = 6.3e-45) in blood. Several phosphatidylcholine metabolites (e.g., PC 32:1 and PC 34:1) were positively associated with alcohol consumption, for example, one g/day higher of alcohol consumption was associated with higher level of PC 32:1 (beta=0.021, p = 3.1e-38) in blood. Among the 20 metabolites (Figure 2) that were negatively associated with alcohol consumption, triacylglycerol 54:4 (TAG 54:4) displayed the most significant association (beta =-0.017, p = 6.17e-22). With additional adjustment for eGFR, the association between alcohol consumption and metabolites remained largely the same (Pearson correlation coefficient=0.99, **Supplemental Figure 1**).

**Table 2.** The Top 20 Significant Metabolites Associated with Cumulative Average Alcohol Consumption.

| Metabolites | Beta | SE | *P* value |
| --- | --- | --- | --- |
| CE 16:1 | 0.023 | 0.002 | 6.30E-45 |
| PC 32:1 | 0.021 | 0.002 | 3.10E-38 |
| PC 34:1 | 0.019 | 0.002 | 5.99E-30 |
| PC-B 36:4 | 0.017 | 0.002 | 5.18E-24 |
| TAG 54:4 | -0.017 | 0.002 | 6.17E-22 |
| PC 32:0 | 0.015 | 0.002 | 1.13E-19 |
| CE 20:5 | 0.014 | 0.002 | 1.14E-17 |
| LPC 20:5 | 0.013 | 0.002 | 2.17E-15 |
| TAG 52:4 | -0.013 | 0.002 | 9.05E-15 |
| TAG 54:5 | -0.013 | 0.002 | 1.18E-14 |
| TAG 52:3 | -0.013 | 0.002 | 1.58E-14 |
| TAG 48:0 | 0.012 | 0.002 | 1.74E-13 |
| fumarate-malate | 0.012 | 0.002 | 7.00E-13 |
| SM 24:1 | 0.012 | 0.002 | 5.17E-12 |
| dimethylglycine | -0.010 | 0.001 | 7.48E-12 |
| xanthurenate | 0.011 | 0.002 | 3.53E-11 |
| PC 34:4 | 0.011 | 0.002 | 9.52E-11 |
| TAG 52:5 | -0.011 | 0.002 | 4.81E-10 |
| PC 36:3 | 0.010 | 0.002 | 7.39E-10 |

Top 20 out of 60 metabolites significantly associated with the cumulative average total alcohol consumption ( $p < 0.00024$ ) adjusting for age, sex, batch, smoking status, BMI, physical activity index and diet score adjusted and family relationship as random effect.

**Figure 2.**
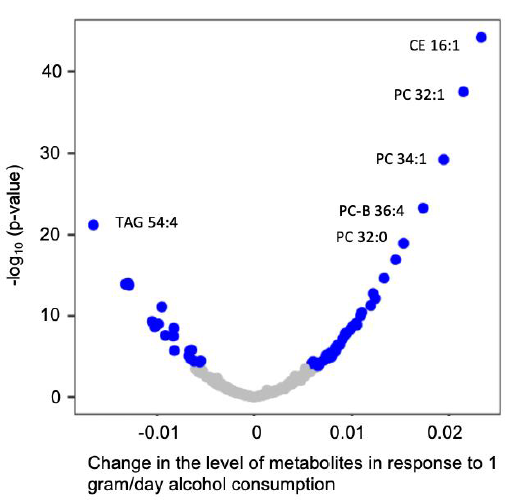
Volcano Plot for Association of Cumulative Average Alcohol Consumption and Metabolites. Linear mixed model was performed adjusting for age, sex, batch, smoking status, BMI, physical activity index and diet score as fixed effect, and family relationship as random effect. Blue dot represents significant relationship (p value < 0.00024), grey dot represents non-significant relationship. CE, cholesteryl ester; PC, phosphatidylcholine; TAG, triacylglycerol.

Roekel’s study found that total alcohol intake was associated with 34 circulating metabolites, including three acylcarnitines, the amino acid citrulline, four lysophosphatidylcholines, 13 diacylphosphatidylcholines, seven acyl-alkylphosphatidylcholines, and six sphingomyelins ^10^. Among the 34 metabolites significant in the Roekel’s study, 13 metabolites were also measured by our metabolite platforms, including LPC 16:0, LPC 16:1, LPC 20:4, PC 32:0, PC 32:1, PC 32:2, PC 34:1, PC 34:3, PC 34:4, PC-B 36:4, PC 38:6, PC 32:1, and SM 24:1. All of the 13 metabolites were significant in our analysis for total alcohol consumption (**Supplemental Table 2**, Supplemental Figure 2).

### Association of each type of alcoholic beverages and metabolites

We found that 19 metabolites were significantly associated with the cumulative average consumption of beer, 30 metabolites were significantly associated with the cumulative average consumption of wine, and 32 metabolites were significantly associated with the cumulative average consumption of liquor (**Supplemental Table 1**, Figure 3). Among the significant ones, seven metabolites (CE 16:1, LPC 20:5, PC 32:0, PC 32:1, PC 34:1, PC-B 36:4, and fumarate-malate) were significantly associated with the cumulative consumption of all three types of alcoholic beverages. (Supplemental Figure 3).

**Figure 3.**
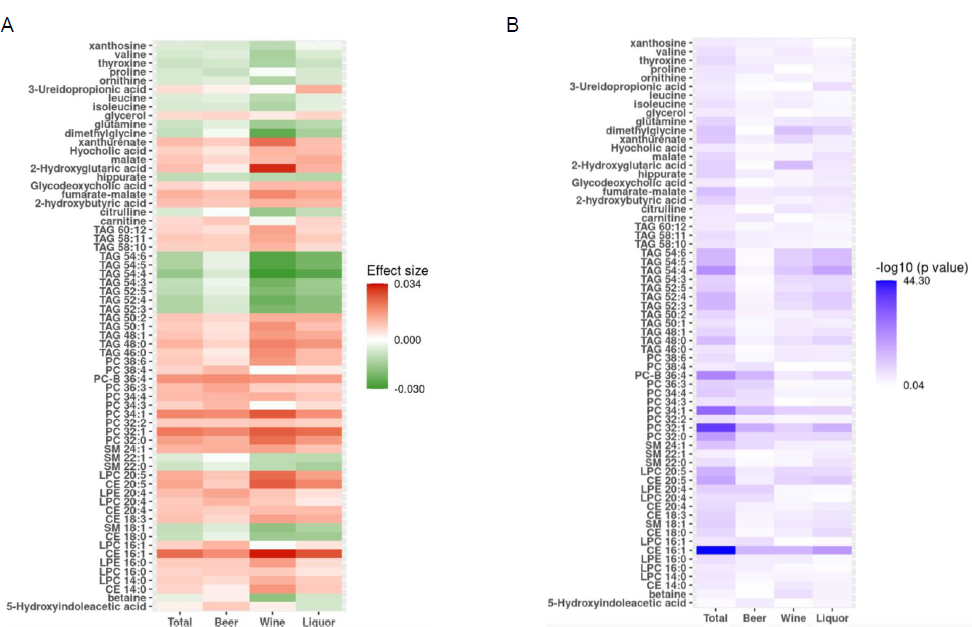
Comparison of Association between Different Types of Alcohol Beverages. Panel A: Comparison of effect size between different types of alcohol beverages; Panel B: Comparison of −log 10 (p value) between different types of alcohol beverages. All linear mixed models adjusted for age, sex, batch, smoking status, BMI, physical activity index and diet score as fixed effect, and family relationship as random effect. CE, cholesterol esters; DAG, diacylglycerols; LPC, lysophosphatidylcholines; LPE, lysophosphatidylethanolamines; PC, phosphatidylcholines; SM, sphingomyelins; TAG, triacylglycerols. Metabolites shown in panel at least had significant association with one type of alcohol (beer, wine and liquor) or total alcohol consumption.

While the three types of alcoholic beverages displayed largely consistent associations with metabolites (Supplemental Figure 4), beer consumption appeared to have slightly weaker associations compared to consumptions of wine and liquor (Supplemental Figure 4). Among 50 metabolites associated with at least one type of alcohol consumption, we calculated the pair-wise ratio of regression coefficients for each metabolite. When we used an absolute ratio of two as cutoff, our analysis revealed that three metabolites (LPC 16:1, PC 34:3 and PC 38:4) displayed stronger association with the cumulative average consumption of beer; one metabolite (3-Ureidopropionic acid) with the consumption of liquor; five metabolites (betaine, 2-hydroxyglutaric acid, xanthurenate, isoleucine and valine) with the consumption of wine (**Supplemental Table 3**).

### Sex-specific associations of metabolite with cumulative average total alcohol consumption

We conducted interaction test for sex and alcohol consumption and found that 13 metabolites (betaine, CE 14:0, CE 16:1, PC 32:0, PC 32:1, PC 34:1, TAG 48:0, TAG 52:4, TAG 54:4, TAG 54:5, xanthurenate, dimethylglycine, and Kynurenic acid) had significant interaction term (p < 0.05/211=0.00024). Eleven (CE 14:0, CE 16:1, PC 32:0, PC 32:1, PC 34:1, TAG 48:0, TAG 52:4, TAG 54:4, TAG 54:5, xanthurenate, and dimethylglycine) of these 13 metabolites were in 60 metabolites that were significantly associated with the cumulative average total alcohol consumption. We applied sex-specific analysis to examine the associations of cumulative average total alcohol consumption with these 13 metabolites. For women, the associations were significant for all of these 13 metabolites (p<0.05/13=0.0038), while for men, 10 metabolites (CE 16:1, PC 32:0, PC 32:1, PC 34:1, TAG 48:0, TAG 52:4, TAG 54:4, TAG 54:5, xanthurenate, and dimethylglycine) had significant associations (**Supplemental Table 4**). The observed sex-alcohol interaction was due to the stronger associations in women than that in men (Figure 4). For example, one g/day higher alcohol consumption was associated with 0.036 standard deviation increases in the level of CE 16:1 in women (p value=7.26e-25), while 0.019 in men (p value=4.25e-22).

**Figure 4.**
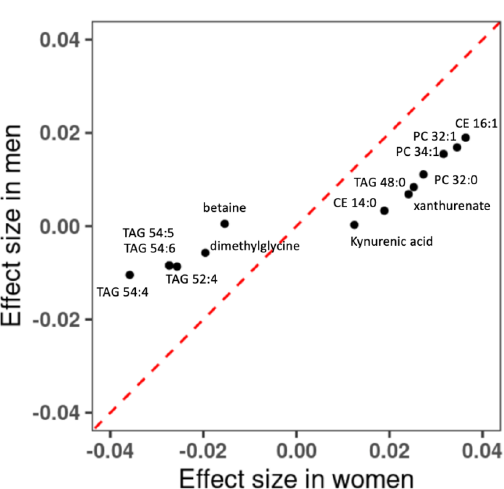
Comparison of Associations of Cumulative Average Alcohol consumption and metabolites between men and women. Linear mixed model was performed in women-only and men-only samples. Covariates included age, batch, smoking status, BMI, physical activity index and diet score as fixed effect, and family relationship as random effect. CE, cholesteryl ester; PC, phosphatidylcholine; TAG, and triacylglycerol.

### Pathway analysis for alcohol-associated metabolites

We conducted pathway analyses of the 60 metabolites that were significantly associated with the cumulative average total alcohol consumption. Using the KEGG library, we found three pathways to demonstrate evidence of enrichment among our significant metabolites? including 1) Arginine biosynthesis (FDR = 0.002); 2) Valine, leucine and isoleucine biosynthesis (FDR =0.04); and 3) Aminoacyl-tRNA biosynthesis (FDR =0.007) (**Supplemental Table 5**). Using the SMPDB library, we did not observe significant pathways at FDR 0.05 level (**Supplemental Table 6)**.

### Association of alcohol-associated metabolites and incident CVD

We included 2138 participants without prevalent CVD in association analyses of the 60 alcohol-associated metabolites with incident CVD. A total of 636 participants developed incident CVD with a median follow-up of 18.1 years. Ten metabolites (TAG 50:2, TAG 50:1, TAG 48:1, TAG 48:0, isoleucine, leucine, TAG 52:3, TAG 46:0, PC 32:1 and glutamine) were significantly associated with the development of CVD (p < 0.05/60≈0.00082) in the base model adjusting for age, sex and batch (**Supplemental Table 7**). Out of the ten metabolites, higher levels of nine metabolites (TAG 50:2, TAG 50:1, TAG 48:1, TAG 48:0, TAG 46:0, PC 32:1, TAG 52:3, isoleucine and leucine) and lower levels of one metabolite (glutamine) were associated with an increased risk of incident CVD (hazard ratio ranges from 1.16 to 1.33, p < 0.05/60≈0.00082). After additionally adjusting for BMI, SBP, hypertension treatment status, diabetes, smoking status, total and high-density lipoprotein cholesterol level, the association with incident CVD remained nominally significant at p <0.05 for four metabolites (TAG 50:2, TAG 50:1, TAG 48:1, and glutamine; **Supplemental Table 7**).

We created two weighted metabolite scores; the first score included six metabolites (TAG 50:2, TAG 50:1, TAG 48:1, TAG 48:0, TAG 46:0 and PC 32:1) that had positive association with both the cumulative average total alcohol drinking and incident CVD in the base models and glutamine, which was negatively associated with both the cumulative average total alcohol drinking and incident CVD in the base model (**Supplemental Table 8**). The second score included three metabolites (TAG 52:3, isoleucine and leucine), which had inverse association with the cumulative average total alcohol drinking and positive association with incident CVD in the base model (**Supplemental Table 8**) In multivariable model analysis, per standard deviation increase of the first metabolite score was associated with 11% higher hazard of incident CVD (p = 0.02, 95%CI=[1.02, 1.21]; Figure 5) and per standard deviation increase of the second metabolite score was associated with 12% lower hazard of incident CVD (p = 0.02, 95%CI=[0.78, 0.98]; Figure 5)

**Figure 5.**
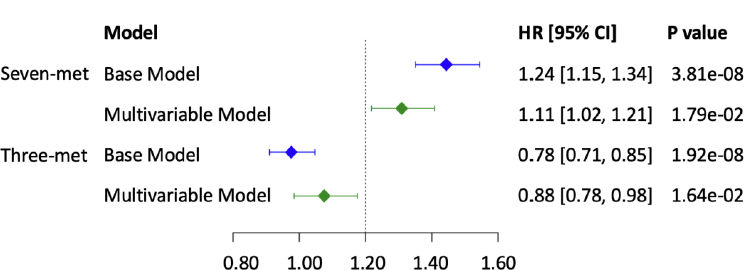
Forest Plot for CVD with Metabolite Composite Score. Seven-met, the weighted score obtained from seven metabolites, TAG 50:2, TAG 50:1, TAG 48:1, TAG 48:0, TAG 46:0, PC 32:1, and glutamine; Three-met, the weighted score obtained from three metabolites, TAG 52:3, isoleucine, and leucine. Weights were obtained from the beta coefficients of association test between cumulative average alcohol consumption and metabolites with Cox regression models. Base model included age, sex, and batch as covariates, and multivariable model additionally adjusted for BMI, SBP, hypertension treatment status, diabetes, smoking status, total and high-density lipoprotein cholesterol level.

## DISCUSSION

In this present study, we investigated the associations between the cumulative average alcohol consumption and 211 circulating metabolites in the 2428 FHS participants. Of the 211 metabolites, the cumulative average alcohol consumption was associated with 60 metabolites. We found that nine metabolites were more strongly associated with a specific type of alcoholic beverage. We also found that the cumulative average total alcohol consumption displayed stronger associations with 13 metabolites in women than men. Furthermore, we created two alcohol consumption-associated metabolite scores and showed that they had comparable but opposite association with incident CVD. Taken together, with targeted metabolomic profiling, our study identified a series of alcohol consumption-associated circulating metabolites, and via these metabolites, alcohol consumption may have counteractive effects on CVD risk.

Our results showed that higher level of alcohol consumption was associated with higher plasma levels for about two third of the 60 significant metabolites. Among the top positively associated metabolites were cholesteryl esters (e.g., CE16:1 and CE20:5), phosphatidylcholine (e.g., PC 32:1) and lysophosphatidylcholine (e.g., LPC 20:5). CEs are lipid molecules that can be hydrolyzed to produce cholesterol and free fatty acids. The accumulation of cholesterol in the arterial intima promotes the development of atherosclerotic plaques in CVD etiology^28^. PC, also called lecithins, is enriched in egg yolk^29^. It is the most abundant phospholipid in cells and positively influences the incorporation of cholesterol in membranes. A previous study in mice found that oral feeding of PC was associated with an increased risk for CVD^30^. LPC is recognized as a key factor associated with cardiovascular diseases^29^.

Fourteen plasma triacylglycerols (TAGs) were among the 60 metabolites that were significant in association analyses with total alcohol consumption. Among these TAGs, six (TAG 52:3, TAG 52:4, TAG 52:5, TAG 54:3, TAG 54:4, and TAG 54:5) displayed negative associations with alcohol consumption (i.e., lower alcohol consumption was associated with higher levels of TAGs) while eight (TAG 46:0, TAG 48:0, TAG 48:1, TAG 50:1, TAG 50:2, TAG 58:10, TAG 58:11, and TAG 60:12) displayed positive associations with alcohol consumption. TAGs emerge as biomarkers of a liver-to-β-cell axis that links hepatic β-oxidation to β-cell functional mass and insulin secretion in pancreas ^31^. TAGs are broken into glycerol and free fatty acids in the process of lipolysis, and free fatty acids are either processed by beta-oxidation or converged to ketone ^32^. In our association analysis for alcohol-associated metabolites and incident CVD, we showed that six TAGs (TAG 50:2, TAG 50:1, TAG 48:1, TAG 48:0, TAG 46:0, and TAG 52:3) were associated with incident CVD using the base models. Among these six TAGs, the association remained significant for TAG 50:2, TAG 50:1, and TAG 48:1 after adjusting for common cardiometabolic CVD risk factors. These observations are in line with the well-known effects of alcohol intake on lipid metabolism^33^.

Alcohol consumption was also associated with several types of circulating metabolites, other than TAGs. For example, we showed that alcohol consumption was associated with reduced levels of dimethylglycine. Dimethylglycine is an amino acid derivative and was associated with CVD risk in Lind’s study^34^. Among the alcohol-associated metabolites, valine, isoleucine, leucine are branched-chain amino acids (BCAA). A recent review summarized the complex relationship between impaired BCAA homeostasis to CVD^35^. Several lines of evidence suggest that higher BCAA levels are associated with increased risk of obesity and diabetes, which is consistent with the present observations on the positive association of leucine and isoleucine with incident CVD. Nonetheless, future studies in larger sample size and diverse populations are needed to examine the relationships between alcohol consumption, BCAA, and CVD.

Our observations also highlight the complex relationship between alcohol consumption and circulating metabolites, which was demonstrated by the analysis using the two metabolite scores. Our observations suggest that, via circulating metabolites, alcohol drinking may have both positive and negative effects on CVD, and the two effects seemed to cancel each other out in our study samples. However, if certain factors disrupt the balance, it is possible that one effect may prevail over the other effect and leads to either increased or decreased risk of developing CVD. As such, future studies are warranted to understand what factors may modify the association of alcohol consumption and circulating metabolites, as well as their impact on the relationship of alcohol consumption with CVD development.

We observed that wine consumption and liquor had stronger associations with TAG, CE and SM lipid metabolites, while beer had stronger associations with PC lipid metabolites. We also found that wine and liquor had different associations with amino acids, quinoline carboxylic acids and hydroxy acids. Liquor consumption was significantly related to higher levels of 3-Ureidopropionic acid. Whereas wine consumption had stronger association with betaine, 2-Hydroxyglutaric acid, xanthurenate, isoleucine and valine compared to liquor. These observations suggest that consumption of different types of alcoholic beverages are associated with different metabolomic responses. Some of these metabolites such as betaine and isoleucine ^35, 36^may play important roles in CVD development. Perhaps due to the short list of metabolites examined, the present study did not support the notion that a certain type of alcohol may bring specific benefits to reduce CVD risk. Future studies including a comprehensive list of metabolites are warranted to investigate this issue.

### Strengths and limitations of the study

Our study had several strengths. The most important advantage was that alcohol drinking data from five exams across around 20 years were utilized in the current study. Our study had several limitations. Despite that the sample size in our study was large, the population was primarily white, middle-aged participants. Therefore, the findings from our study may not be generalizable to populations of different races and age groups. Alcohol consumption was measured by questionnaires and calculated based on standard serving size. This approach is cost-effective; however, measurement errors may bias the observed association between alcohol intake and metabolite levels. We acknowledge that the metabolites data analyzed in our study are comprised of a targeted panel. Association between alcohol drinking and untargeted metabolites remained to be studied. We observed that sex may modify the association between total alcohol consumption and metabolite levels. Several other factors may also modify the observed associations. For example, presence or absence of food in the stomach can change the rate of alcohol absorption and metabolism ^37, 38^, and subsequently affect the association of alcohol consumption with circulating metabolite profiles. However, the information about when participants consumed alcohol, for example before or after meals, was not collected. Participants’ genetic background may be a key factor^39^, however, we did not attempt to study that in the present analysis given that analysis may need much larger sample size.

In conclusion, the present study identified a series of alcohol-associated circulating metabolites. Our observations suggest that, via some metabolites, alcohol consumption may have counteractive effects on CVD. Future studies are required to validate our findings and to investigate factors that may modify the associations between alcohol consumption and metabolite levels, particularly for metabolites that potentially contribute to CVD risk, in larger and diverse study samples.

## ABBREVIATIONS

CVD: cardiovascular disease
CE: cholesterol ester
DASH: Dietary Approaches to Stop Hypertension
DAG: diacylglycerol
eGFR: estimated glomerular filtration rate
FDR: false discovery rate
FHS: Framingham Heart Study
HR: hazard ratio
KEGG: Kyoto Encyclopaedia of Genes and Genomes
LC/MS: liquid chromatography with tandem mass spectrometry
LPC: lysophosphatidylcholine
LPE: lysophosphatidylethanolamine
PC: phosphatidylcholine
SBP: Systolic blood pressure
SD: standard deviation
SE: standard error
SM: syphingomyelin
SMPDB: Small Molecule Pathway Database
TAG: triacylglycerol

## DECLARATIONS

### Ethics approval and consent to participate

All participants provided written informed consent. All FHS study protocols were approved by the Boston University Medical Center Institutional Review Board.

### Consent for publication

All authors approve publication of the final manuscript.

### Availability of data and materials

The Framingham Heart Study datasets analyzed in the present study are available at the dbGAP Study Accession: phs000007.v33.p14 (https://www.ncbi.nlm.nih.gov/projects/gap/cgi-bin/study.cgi?study_id=phs000007.v33.p14) via the Controlled Access Data.

### Disclaimer

The views and opinions expressed in this manuscript are those of the authors and do not necessarily represent the views of the National Heart, Lung, and Blood Institute, the National Institutes of Health, or the U.S. Department of Health and Human Services.

### Author contributions

Conceptualization: CL, JM

Methodology: CL, JM, YL

Investigation: CL, JM, YL

Visualization: CL, JM, YL

Funding acquisition: CL, JM

Project administration: CL, JM

Supervision: CL, JM

Writing – original draft: YL, CL, JM

Writing – review & editing: YL, MW, XL, JR, PEM, RJ, TH, XG, JR, JS, BY, MN, DL, CL, JM

## Acknowledgements

The authors thank the staff and participants of the Framingham Heart Study for their important contributions.

---

**Supplemental Figure 1.**
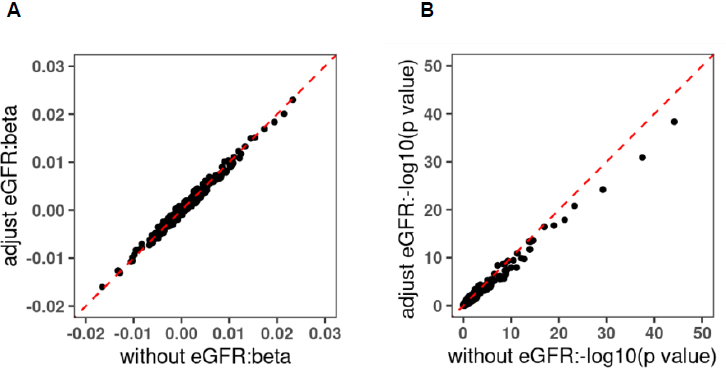
Comparison of Association of Alcohol Consumption and Metabolites with or without Adjusting for eGFR. Panel A: comparison of beta; Panel B: comparison of −log10 (p value). Two models both were performed adjusting for age, sex, batch, smoking status, BMI, physical activity index and diet score, and family relationship as random effect. eGFR, estimated glomerular filtration rate.

**Supplemental Figure 2.**
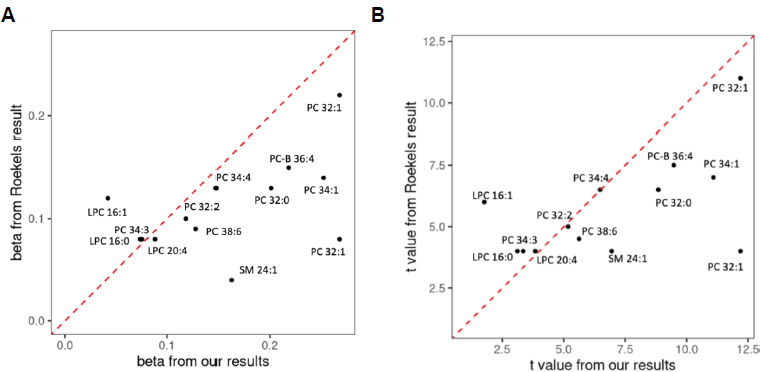
Comparison with Roekel’s Study. Panel A: comparison of beta; Panel B: comparison of t value. Only in this analysis, alcohol consumption (g/day) in this study was plus 1 and natural log transformed. The sex and batch-adjusted residual of metabolites from linear mixed model were used as outcome. Then we applied linear mixed model for alcohol drinking and residual of metabolites in linear mixed model, adjusting for age, sex, smoking status, BMI, physical activity, diet score as fixed effect, and familial relationship as random effect. But Roekel’s study used linear model and covariates included age at blood collection, sex, country, fasting status at blood collection, smoking status, BMI, Cambridge physical activity index, and daily intake of energy, meat and meat products, fish, and shellfish.

**Supplemental Figure 3.**
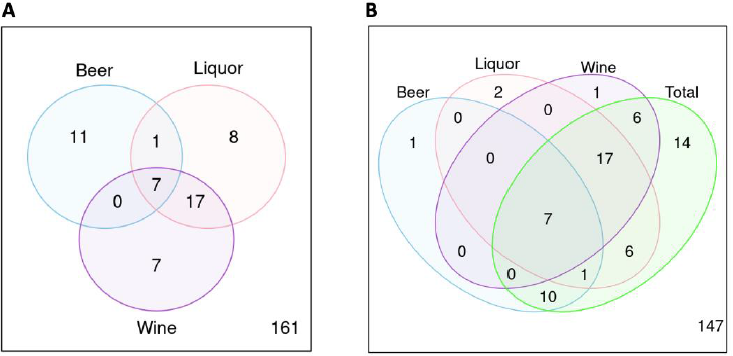
Number of Metabolites Significantly Associated with Each Type of Alcohol Consumption. Panel A: three type of alcohol consumption; Panel B: three type of alcohol consumption and total alcohol consumption. All models were performed adjusting for age, sex, batch, smoking status, BMI, physical activity index and diet score, and family relationship as random effect.

**Supplemental Figure 4.**
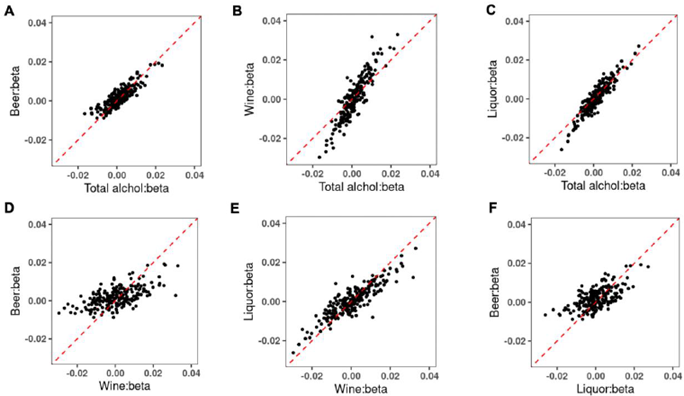
Comparison of Association Analyses of Alcohol Consumption with Metabolites. Panel A-C, comparison of effect size from association analyses of cumulative average total alcohol consumption and each type of alcohol consumption with metabolites. Panel D-F, comparison of effect size from association analyses each type of alcohol consumption with metabolites. All models were performed adjusting for age, sex, batch, smoking status, BMI, physical activity index and diet score, and family relationship as random effect.

